# Clinical Characteristics and Patterns of Valve Lesions in Rheumatic Heart Disease Among Children at Hiwot Fana Comprehensive Specialized Hospital: A Comparative Study of Newly Diagnosed and Known Cases

**DOI:** 10.1101/2024.05.08.24307081

**Authors:** Temesgen Teferi Libe, Yunus Edris Kelil, Samrawit Abebaw Tegene, Faisal Abdi, Kibrom Hailemariam Mesfin

## Abstract

**Background:** Rheumatic heart disease remains a significant health burden in resource-limited settings. This study investigates the clinical characteristics and valve lesion patterns of RHD in children from Eastern Ethiopia, comparing newly diagnosed and known RHD patients.

**Objective:** This study aims to characterize the clinical features and valve lesion patterns in children with Rheumatic heart disease and provide a comparative analysis between newly diagnosed and known cases.

**Methods:** A hospital-based cross-sectional study was conducted at Hiwot Fana Comprehensive Specialized Hospital from January 1 to December 31, 2021. A total of 39 children with RHD were included, with data collected from medical records, clinical assessments, and echocardiographic evaluations. Descriptive statistics and chi-square tests were used for analysis.

**Results:** Among the 39 children studied, 25 were newly diagnosed and 14 were known RHD cases. The majority were female (71.8%). The median age was 10 years. Shortness of breath (53.9%) and cough (38.5%) were the most common presenting complaints. Only 14.3% of known RHD patients were adherent to secondary prophylaxis. Severe acute malnutrition and severe anemia were the most common comorbidities. Class IV heart failure was present in 89.7% of the patients. Echocardiographic findings revealed that all patients had mitral valve involvement, with mitral regurgitation (94.9%) being the most frequent.

**Conclusion:** The study highlights significant clinical characteristics and valve lesion patterns among children with Rheumatic heart disease at Hiwot Fana Comprehensive Specialized Hospital, emphasizing the need for early diagnosis, improved adherence to prophylaxis, and targeted interventions to manage comorbidities and advanced heart failure.

## Introduction

Rheumatic heart disease (RHD) remains a significant public health threat, particularly for children in developing countries like Ethiopia. It is a debilitating condition primarily affecting the heart valves, particularly the mitral and aortic valves, resulting from acute rheumatic fever (ARF), a complication of untreated streptococcal pharyngitis[1]. The global burden of RHD is immense, with developing nations disproportionately affected due to factors like poverty, overcrowding, and limited access to healthcare[2–9]. In children, RHD can hinder growth and development, making early diagnosis and intervention crucial[10].

Despite the substantial national burden of RHD, particularly in Ethiopia’s Eastern region, there is a scarcity of studies exploring the clinical characteristics and patterns of valve lesions in children. This study aims to address this critical knowledge gap. The lack of understanding concerning the specific clinical features and valve lesion patterns in this population hinders the development of tailored interventions, early detection, and optimal management of RHD.

Furthermore, a comparative analysis of the clinical characteristics and patterns of valve lesions in newly diagnosed and known RHD groups will help identify potential gaps in the prevention and management of the disease, whether due to delayed diagnosis or subsequent management issues. Therefore, this study aimed to comprehensively characterize the clinical features and valve lesion patterns in children with RHD and provide a comparative analysis of these conditions in newly diagnosed and known cases.

## Methods and Materials

### Study area, design and period

A hospital-based cross-sectional study design was used to prospectively study children admitted from January 1 to December 31, 2021, to the pediatric ward and pediatrics ICU of Hiwot Fana Comprehensive Specialized Hospital (HFCSH) who were diagnosed with RHD. HFCSH, Haramaya University’s tertiary teaching hospital, is located in Harar and serves 5.8 million people, including the populations of the Harari Region, Diredawa City Administration, Somali Region, and the Eastern Hararghe Zone of Oromia Regional State. At HFCSH, the pediatric ward and ICU accommodate 30 beds (6 in PICU and 24 in the pediatric ward) with about 640 total admissions per year, among which approximately 72 had an RHD diagnosis (Health Management Information System data, 2019; unpublished).

### Patient involvement

Patients were not directly involved in the design of this study.

### Population

Thirty-nine children with RHD admitted to the pediatric ward and pediatric ICU of HFCSH from January 1 to December 31, 2021, were studied after excluding 21 repeat admissions and 7 disappeared cases before obtaining adequate information. (Fig 1)

**Figure 1:**
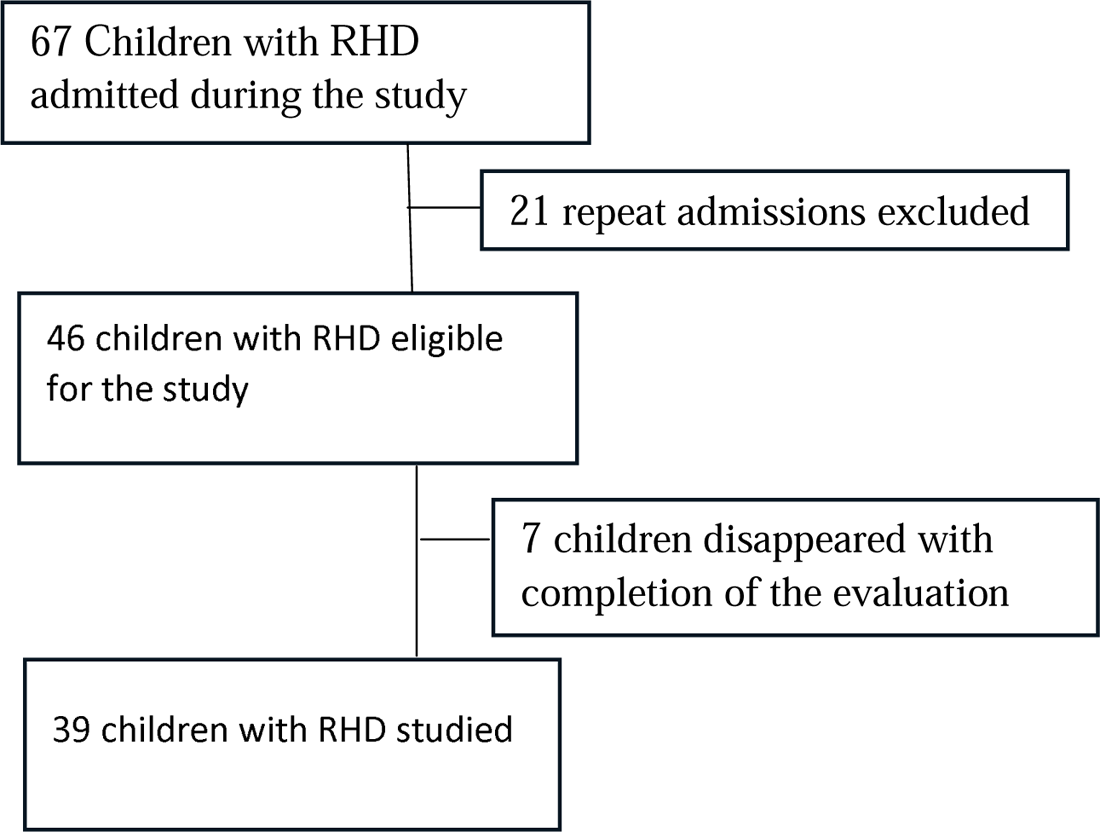
Patient selection procedure

### Data Collection Methods

A comprehensive review of medical records for children with rheumatic heart disease (RHD) admitted to the pediatric ward and ICU at Hiwot Fana Comprehensive Tertiary Teaching Hospital between January 1 and December 31, 2021, was conducted. The review included demographic information, clinical history, presenting symptoms, laboratory results, chest X-ray findings, and echocardiography reports.

An experienced team of pediatricians and cardiologists conducted standardized clinical assessments on each enrolled child (n= 39) from January 1 to December 31, 2021. These assessments included a thorough physical examination for cardiac symptom evaluation, vital sign measurement, and collection of additional clinical data.

Pediatric cardiologists performed echocardiographic assessments adhering to standardized protocols to identify valve lesions and their characteristics during the same period (January 1 – December 31, 2021). Additionally, laboratory data (complete blood count, ESR, and creatinine), chest X-ray, and ECG were obtained.

### Data quality control

The data collection tool was piloted on two RHD patients in the Jugol Hospital pediatric ward to assess its consistency, completeness, and ease of understanding. Two days of training were given to both the data collectors and supervisor. The principal investigator closely monitored the data collection process to ensure completeness, accuracy, and data collection consistency. During sessions, thorough checking was done before analyzing the filled questionnaires. Two separate individuals conducted double data entry. When any gap was identified, it was communicated with data collectors daily. Following data entry, the dataset was reviewed for completeness and any missing data points were addressed.

### Data Processing and Analysis

After data collection, each questionnaire was checked thoroughly for completeness. The data was then coded using a standardized coding scheme and double-entered into EpiData version 3.1. Subsequently, the data was transferred to Stata version 14.2 for statistical analysis.

Descriptive statistics, including frequency, proportions, means, and standard deviation were computed. Chi-square tests were used to identify any association between RHD status and dependent variables. Due to the small sample size, Fisher’s exact method was also employed.

### Ethical consideration

To ensure ethical conduct throughout the research process, approval was obtained from the Haramaya University College of Health and Medical Sciences Institutional Health Research Ethical Review Committee (reference number: IHRERC/245/2020). The study was conducted in strict accordance with the principles outlined in the Declaration of Helsinki.

Following the ethical approval process, written permission for data access was obtained from the hospital managers/medical directors of Hiwot Fana Comprehensive Specialized Hospital. To safeguard patient privacy, the study utilized anonymized data throughout the analysis. This approach ensured that researchers did not have access to any personally identifiable information (PII) such as medical record numbers, phone numbers, or patient names. Since data was collected from medical charts, consent from parents/guardian was not needed.

## Result

### Baseline Characteristics

During the study period, a total of 67 children with RHD were admitted. However, 21 were repeat admissions, and 7 disappeared before completing the evaluation. Therefore, 39 children with RHD were enrolled in the study. [Fig 1]

Fourteen children had a known diagnosis of RHD, and 25 were newly diagnosed. The majority (71.8%, n=28) were females, while the remaining participants (28.2%, n=11) were males. Among the 25 newly diagnosed cases, 18 (72%) were female and 7 (28%) were male. Among the 14 known RHD cases, 10 (71.4%) were female and 4 (28.6%) were male. [Fig 2]

**Figure 2:**
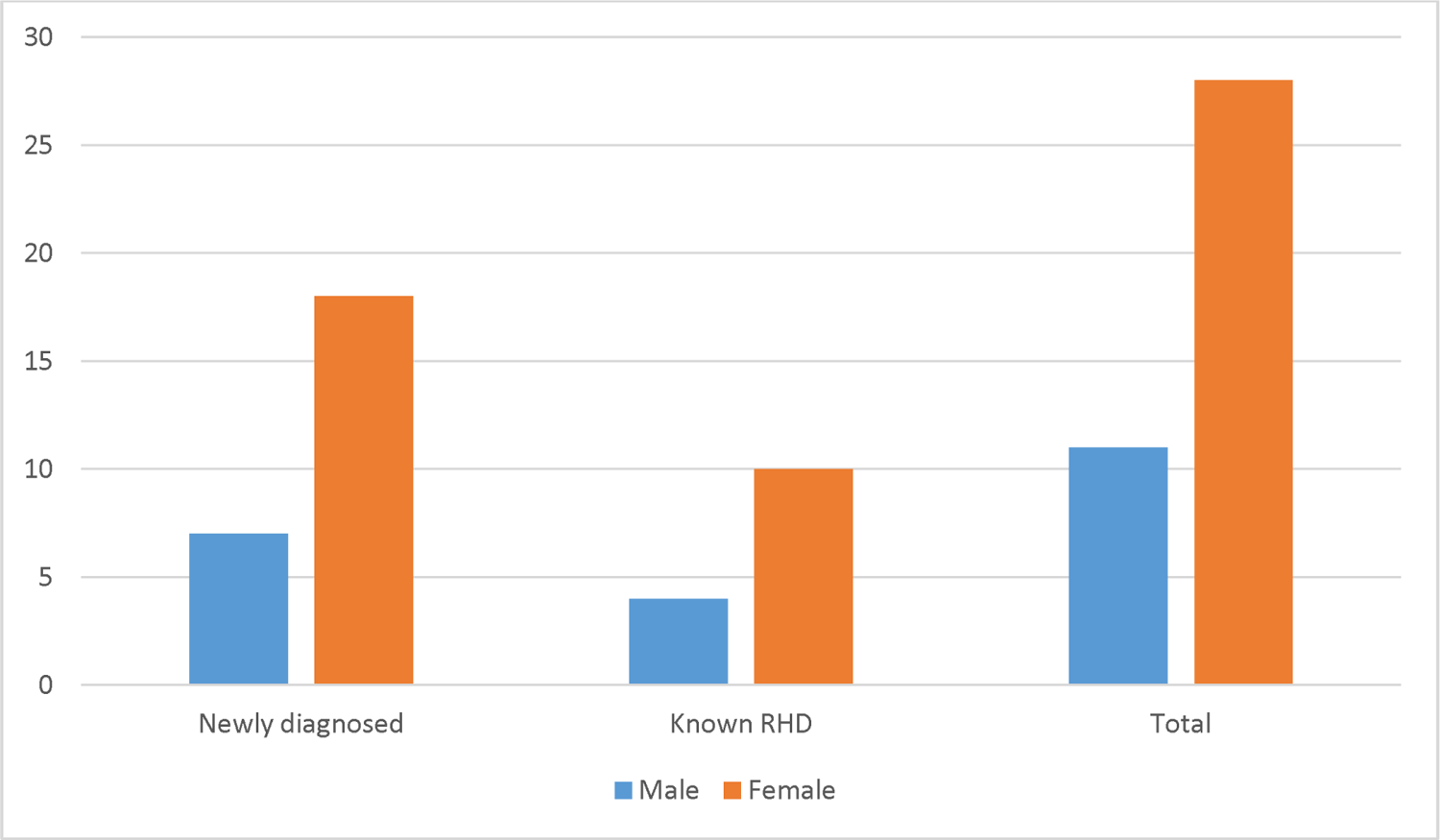
Sex of children admitted with diagnosis of RHD at HFCSH

The median age of the children was 10 years with an IQR of 4. In newly diagnosed RHD cases, the median age was 10 with an IQR of 5, while in known RHD cases, the median age was 10.5 with an IQR of 4. There is no statistically significant difference in the mean age between children with newly diagnosed RHD and those with known RHD, as indicated by the t-test (two-tailed p-value = 0.29).

### Clinical characteristics

Shortness of breath (53.9%) was the most frequent presenting complaint, followed by cough (38.5%). When comparing presenting complaints between new and known RHD patients, 12 (48%) of the newly diagnosed RHD patients presented with shortness of breath, and 10 (40%) presented with a cough. In the known RHD patient group, 9 (64.2%) presented with shortness of breath, and 5 (35.7%) presented with a cough. (Table 1)

**Table 1:**
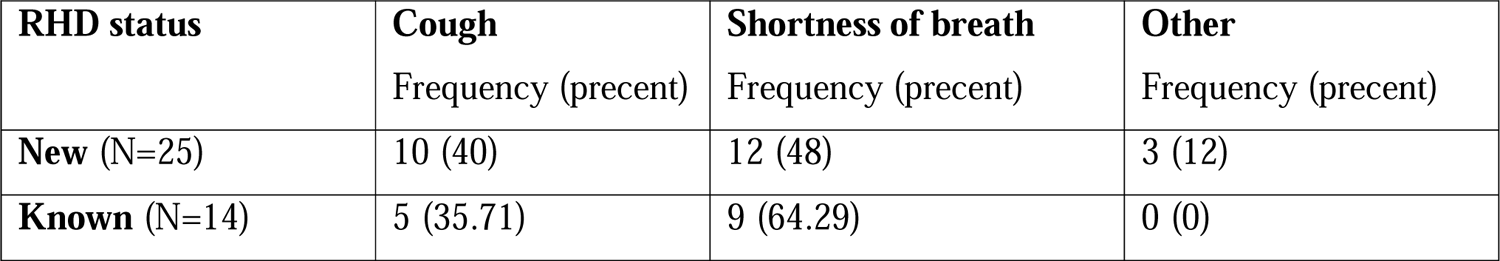
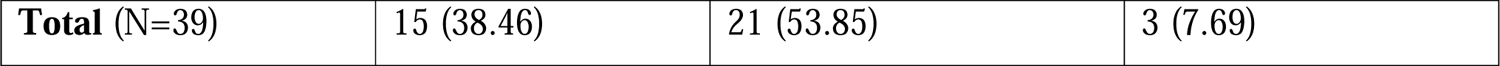
Presenting complaint of children diagnosed with RHD at HFCSH.

Among the 14 children with a known diagnosis of RHD, only 2 (14.3%) were adherent to secondary prophylaxis, while 6 (42.9%) were non-adherent, and the remaining 6 (42.9%) had never taken prophylaxis. (Fig 3)

**Figure 3:**
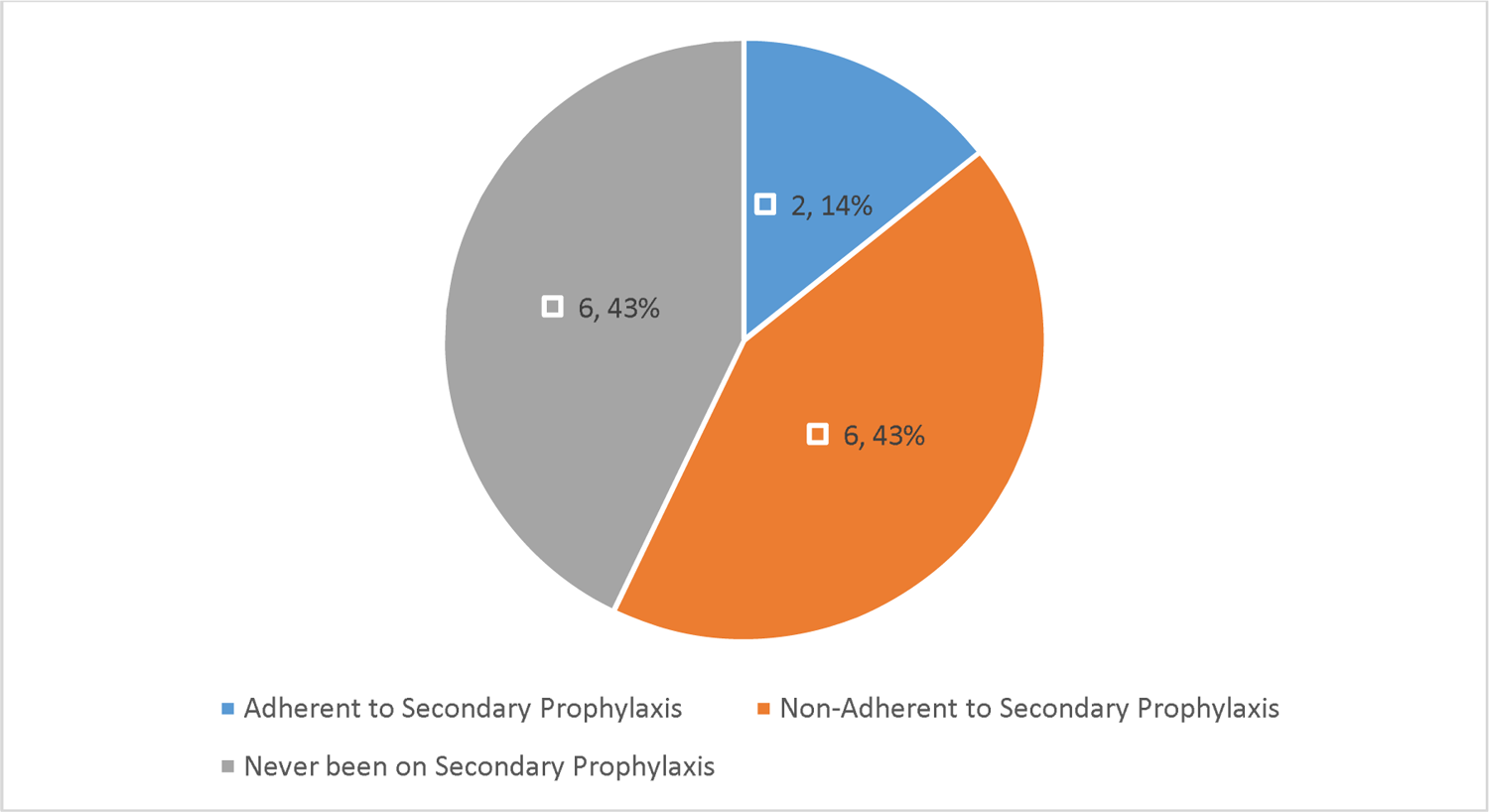
Adherence of known RHD patients at HFCSH to secondary prophylaxis.

Ten (25.6%) children had at least one comorbidity. The most frequent comorbidities were severe acute malnutrition (5; 12.8%) and severe anemia (Hb < 7 gm/dl; 5; 12.8%). Among the newly diagnosed group, 7 had comorbidities, with 3 cases of severe acute malnutrition and 4 cases of severe anemia. Among the 14 known RHD group, 3 had comorbidities, including 2 cases of severe acute malnutrition and 1 case of severe anemia. (Fig 4)

**Figure 4:**
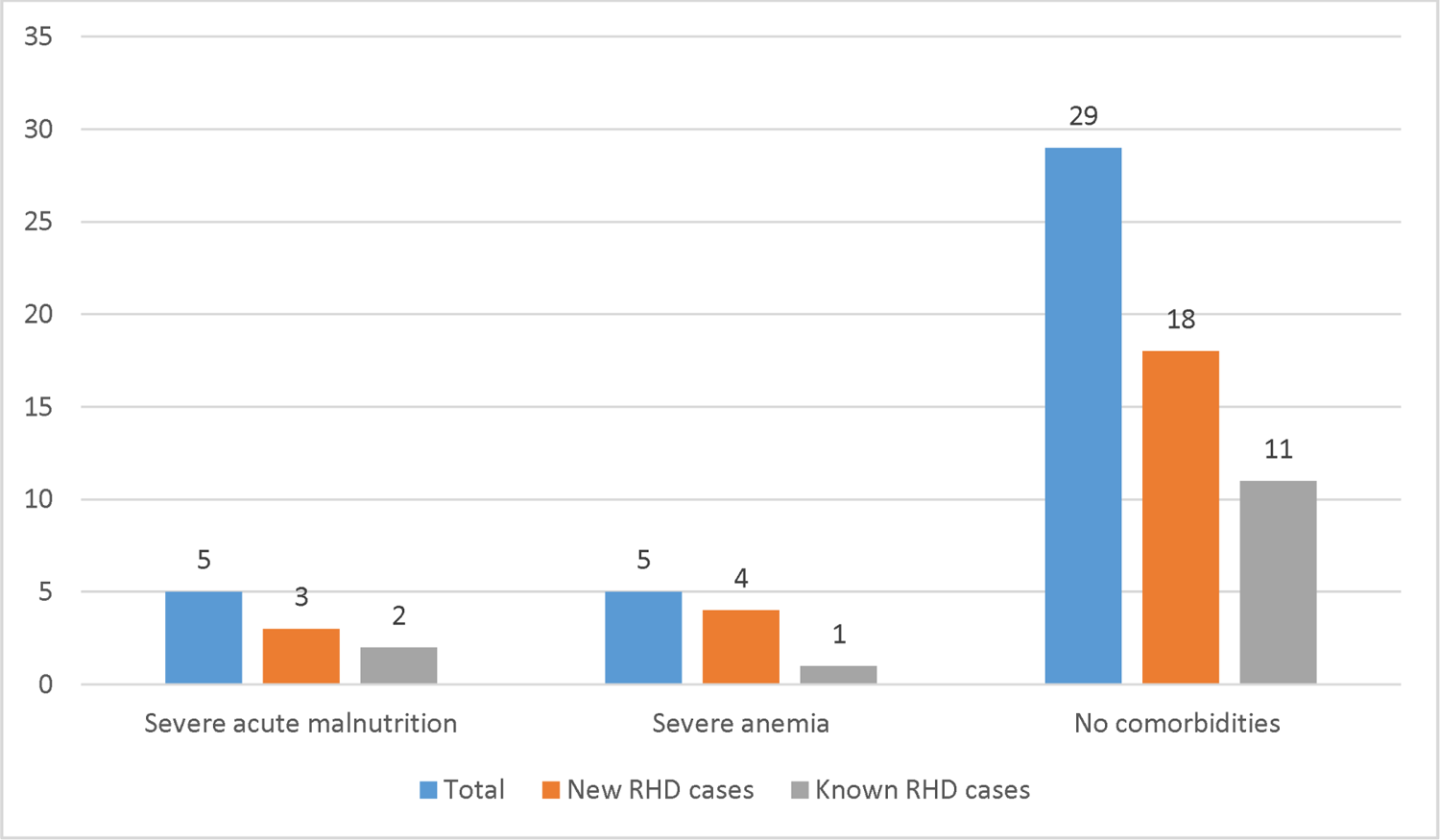
Comorbidities in Children with RHD admitted to HFSCH.

A significant proportion of patients (35 out of 39, or 89.7%) presented with Class IV Heart Failure upon admission. Among these 35 patients, 23 were newly diagnosed cases of RHD, while 12 were known RHD cases. (Table 2)

**Table 2:** Class IV Heart Failure Up on Admission.

| RHD status | Class IV Heart Failure |
| --- | --- |
| New (N=25) | 23 (92) |
| Known (N=14) | 12 (85.7) |
| Total (N=39) | 35 (89.7) |

Among patients admitted with class IV heart failure, pneumonia was the most frequently identified trigger, affecting 20% (7 patients). Rheumatic fever was identified in 11% (4 patients), and infective endocarditis was confirmed or suspected in 14% (5 patients). (Table 3)

**Table 3:**
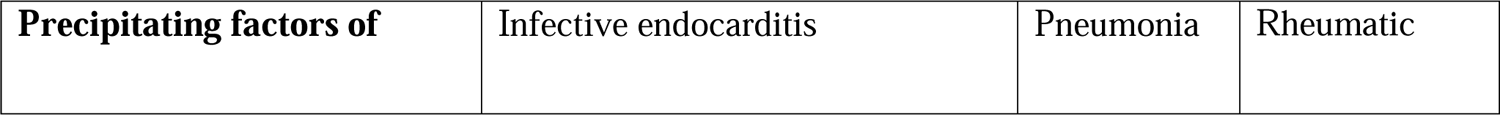
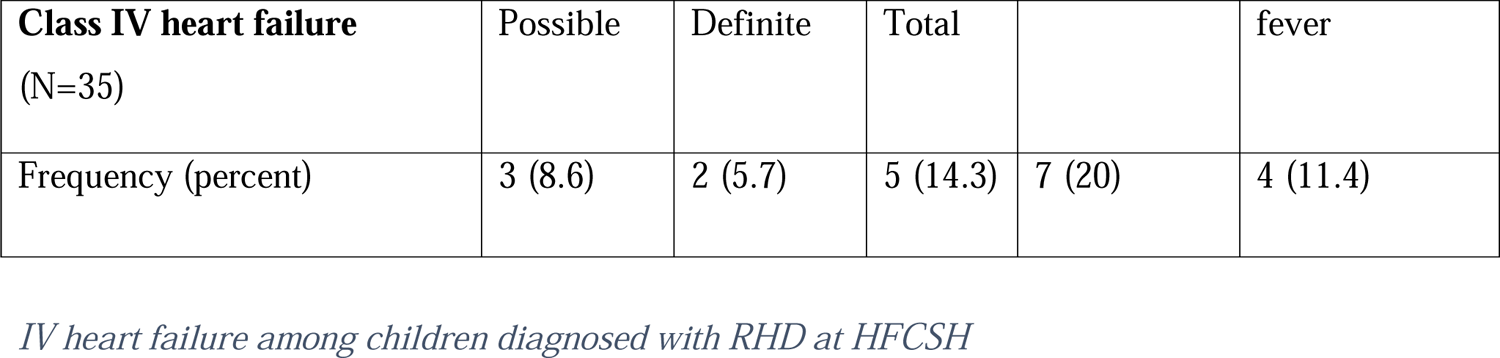
Precipita ting factors of class.

| Precipitating factors of | Infective endocarditis | Pneumonia | Rheumatic |
| --- | --- | --- | --- |

|  |  |  |  |  |  |
| --- | --- | --- | --- | --- | --- |
| **Class IV heart failure**<br>(N=35) | Possible | Definite | Total |  | fever |
| Frequency (percent) | 3 (8.6) | 2 (5.7) | 5 (14.3) | 7 (20) | 4 (11.4) |

Of the 4 (10.3%) rheumatic fever cases among 39 RHD patients, 1 (4% of the group) occurred in the Newly diagnosed RHD patients, whereas 3 (21.4% of the group) occurred in the Known RHD patients. (Table 4)

**Table 4.**
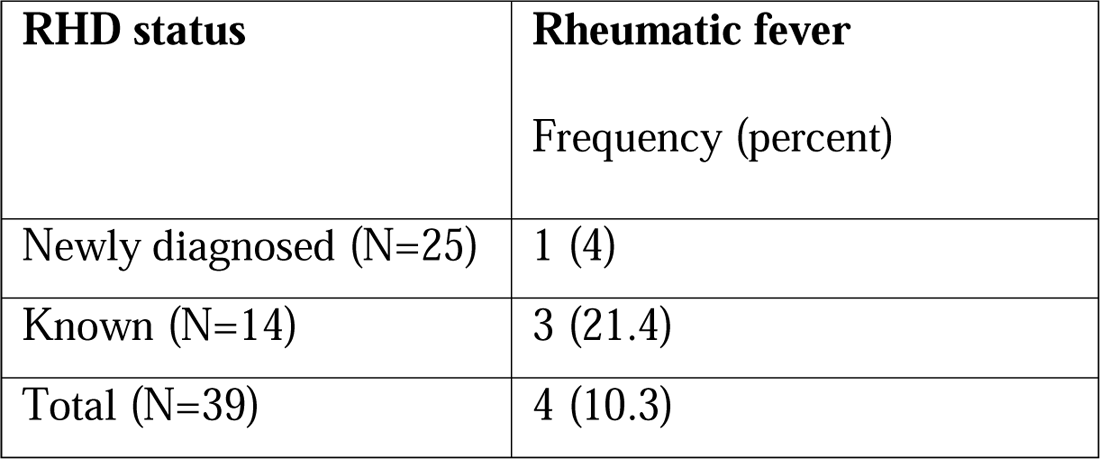
Rheumatic fever among RHD patients at HFCSH.

| **RHD status** | **Rheumatic fever**<br>Frequency (percent) |
| --- | --- |
| Newly diagnosed (N=25) | 1 (4) |
| Known (N=14) | 3 (21.4) |
| Total (N=39) | 4 (10.3) |

### Vital signs and Anthropometry

The median pulse rate was 116 beats per minute (bpm) with an IQR of 21 bpm. In the newly diagnosed RHD group, the median pulse rate was 115 bpm with an IQR of 24 bpm, whereas in the known RHD group, the median pulse rate was 120 bpm with an IQR of 16 bpm.

The median respiratory rate was 36 breaths per minute with an IQR of 22 breaths per minute. In the newly diagnosed RHD group, the median respiratory rate was 36 breaths per minute with an IQR of 18 breaths per minute, whereas in the known RHD group, the median respiratory rate was 36 breaths per minute with an IQR of 24 breaths per minute.

The median body temperature was 36.9°C with an IQR of 0.9°C. In the newly diagnosed RHD group, the median temperature was 36.9°C with an IQR of 0.4°C, whereas in the known RHD group, the median temperature was 36.95°C with an IQR of 1.3°C.

The median oxygen saturation on room air was 95.5% with an IQR of 5%. In the newly diagnosed RHD group, the median oxygen saturation was 96% with an IQR of 5.5%, whereas in the known RHD group, the median was 93.5% with an IQR of 5%.

The median body mass index (BMI) was 13.85 kg/m² with an IQR of 2.9 kg/m². In the newly diagnosed RHD group, the median BMI was 14 kg/m² with an IQR of 2.7 kg/m², whereas in the known RHD group, the median BMI was 12.9 kg/m² with an IQR of 2.6 kg/m². (Table 5)

**Table 5:**
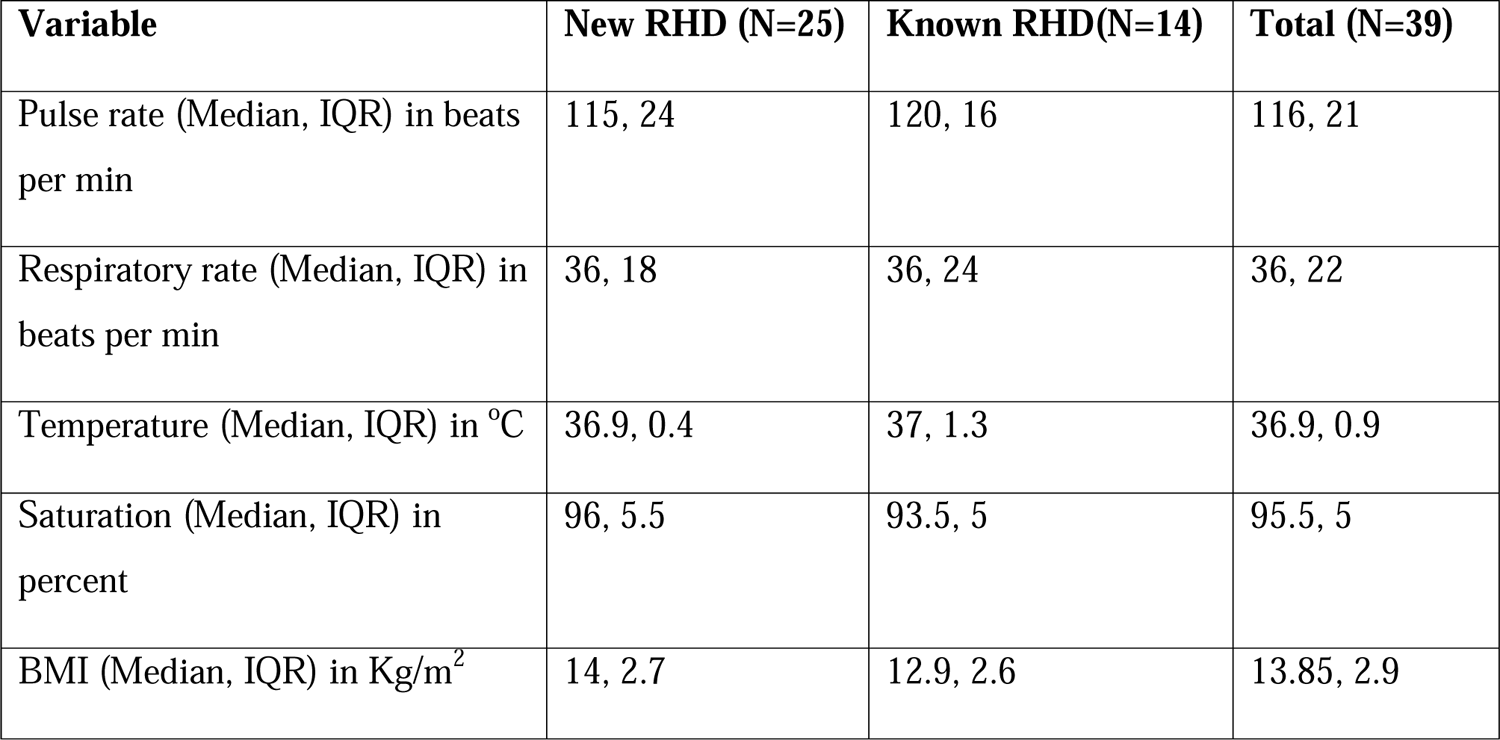
Vital sign and BMI distribution of Children with RHD admitted to HFCSH.

According to WHO Z-scores for age, 5 (12.8%) children were severely wasted (< - 3 Z-score), 12 (30.8%) were moderately wasted (−3 to −2 Z-score), 21 (53.8%) had normal weight (−2 to +2 Z-score), and 1 (2.6%) was overweight (> +2 Z-score). (Figure 5)

**Figure 5:**
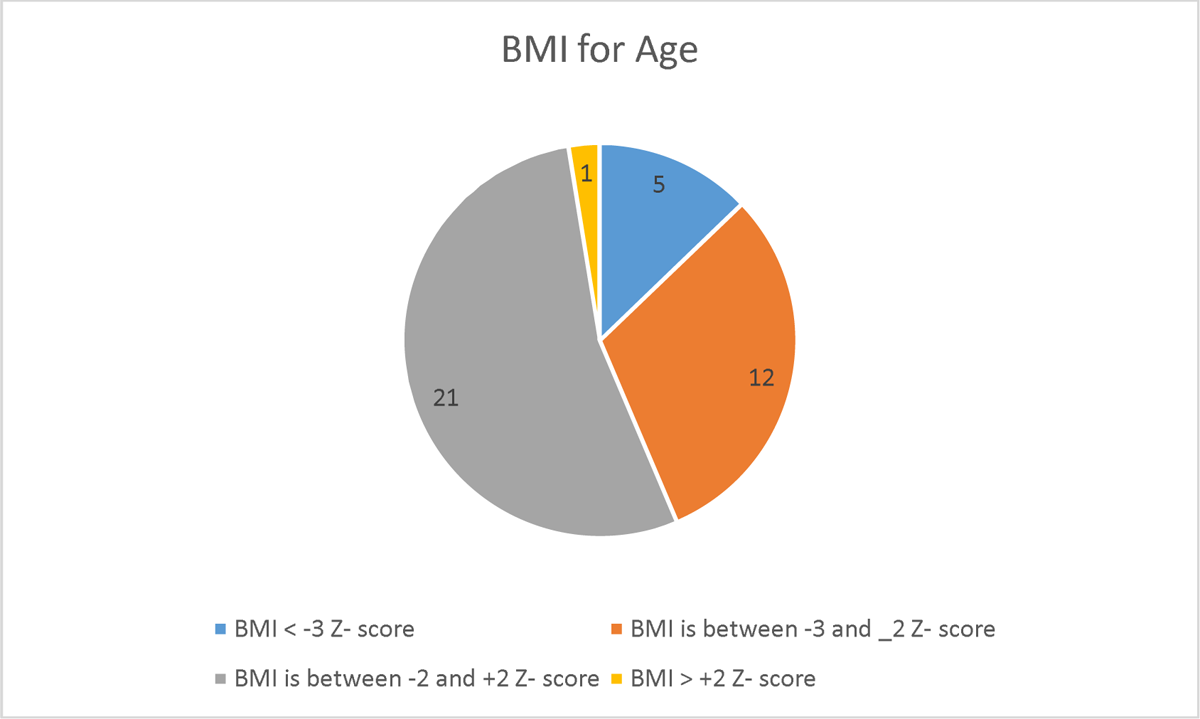
BMI for Age distribution of Children with RHD admitted to HFCSH.

### Chest examination

At least one abnormal chest examination finding was present in 24 (61.5%) children. The most common findings were crepitations (41.0%), decreased or absent air entry (39.5%), dullness (39.5%), and wheezing (9.3%). (Table 6)

**Table 6:**
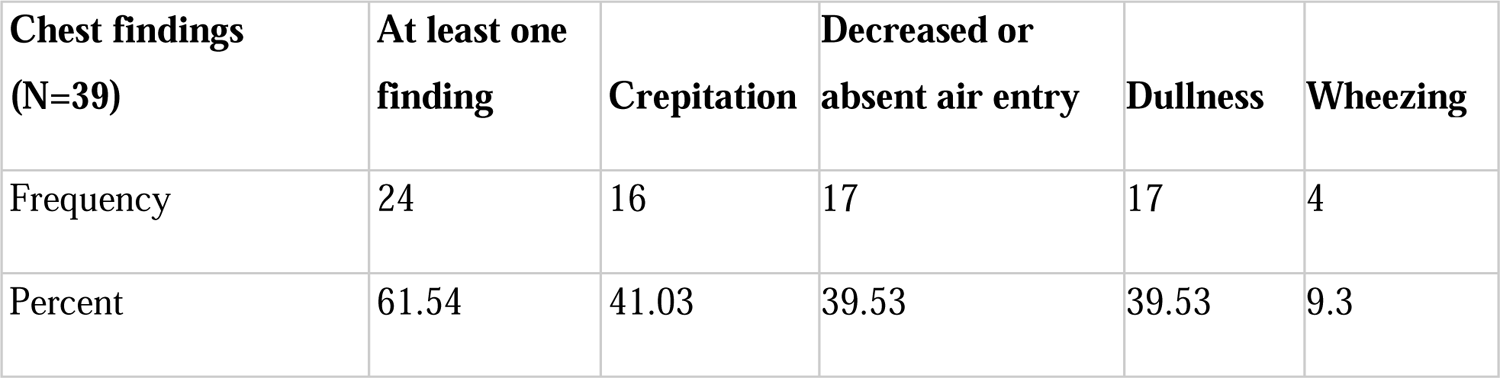
Chest examination findings of Children with RHD admitted to HFCSH.

| **Chest findings**<br>**(N=39)** | **At least one**<br>**finding** | **Crepitation** | **Decreased or**<br>**absent air entry** | **Dullness** | **Wheezing** |
| --- | --- | --- | --- | --- | --- |
| Frequency | 24 | 16 | 17 | 17 | 4 |
| Percent | 61.54 | 41.03 | 39.53 | 39.53 | 9.3 |

### Cardiac examination

The point of maximal impulse (PMI) was displaced downward and to the left in 28 (71.8%) children, while it was in the normal position in 9 (23.1%) and on the left lower sternal border in 2 (5.1%). Apical heave (61.5%) and parasternal heave (56.4%) were the most frequent abnormal cardiac examination findings. P2 heart sound was accentuated in 28 (77.8%) children. A third heart sound (S3 gallop) was present in 5 (12.8%) children. Almost all children (97.4%) had a significant murmur on cardiac auscultation. (Table 7)

**Table 7:** Cardiac examination findings of Children with RHD admitted to HFCSH.

| **Point of Maximum impulse (N=39)** | **Freq. (Percent)** |
| --- | --- |
| Normal position | 9 (23.08) |
| On left lower sternal | 2 (5.13) |
| Deviated to left and/or downward | 28 (71.79) |
| Total | 39 (100) |
| **Apical Heave (N=39)** | **Freq. (Percent)** |
| Yes | 24 (61.54) |
| No | 15 (38.46) |
| Total | 39 (100) |
| **Parasternal heave (N=39)** | **Freq. (Percent)** |
| Yes | 22 (56.41) |
| No | 17 (43.59) |
| Total | 39 (100) |
| **S3 gallop (N=39)** | **Freq. (Percent)** |
| Yes | 5 (12.82) |
| No | 34 (87.18) |
| Total | 39 (100) |

### Laboratory findings

The median hemoglobin (Hb) concentration was 10.3 g/dL with an IQR of 3.5 g/dL. In the newly diagnosed RHD group, the median Hb concentration was 10.2 g/dL with an IQR of 2.8, whereas in the known RHD group, the median Hb concentration was 11.35 g/dL with an IQR of 4.

The median erythrocyte sedimentation rate (ESR) was 40 mm/hr with an IQR of 40 mm/hr. In the newly diagnosed RHD group, the median ESR was 35 mm/hr with an IQR of 35 mm/hr, whereas in the known RHD group, the median ESR was 45 mm/hr with an IQR of 35 mm/hr.

The median serum creatinine level was 0.4 mg/dL with an IQR of 0.2 mg/dL. In the newly diagnosed RHD group, the median serum creatinine level was 0.4 mg/dL with an IQR of 0.1 mg/dL, whereas in the known RHD group, the median was 0.4 mg/dL with an IQR of 0.6 mg/dl.

### Chest x ray findings

Chest X-ray revealed abnormal findings in a significant proportion of participants. Left atrial enlargement was the most frequent abnormality, observed in 86.1% (31) of patients. Other findings included cardiomegaly (80.6%), pulmonary edema (26.5%), signs suggestive of pulmonary arterial hypertension (PAH; 36.7%), and pleural effusion (24.2%). (Table 8)

**Table 8.** Chest X-Ray Findings of Children with RHD to HFCSH.

| **Chest X-Ray Finding (N=36)** | **Frequency (percent)** |
| --- | --- |
| Left Atrial Enlargement | 31 (86.1) |
| Cardiomegaly | 29 (80.6) |
| Pulmonary Edema | 9 (26.5) |
| Pulmonary Arterial Hypertension | 11 (36.7) |
| Pleural Effusion | 8 (24.2) |
| Pneumonia | 7 (21.2) |

### Echocardiographic Findings in RHD Patients

Echocardiography revealed significant valvular involvement in a vast majority of patients with RHD. All 39 patients showed evidence of mitral valve involvement. The most frequent finding was mitral regurgitation (MR), present in 37 patients (94.9%). MR was found in 23 patients (92%) in the New RHD group and all 14 patients (100%) in the Known RHD group. The severity of MR varied: 3 patients (7.7%) had mild MR, 7 patients (17.9%) had moderate MR, and 27 patients (69.2%) had severe MR. Specifically, in the New RHD group, 2 patients (8%) had mild MR, 5 patients (20%) had moderate MR, and 16 patients (64%) had severe MR; whereas in the Known RHD group, 1 patient (7.14%) had mild MR, 2 patients (14.28%) had moderate MR, and 11 patients (78.57%) had severe MR.

Mitral stenosis (MS) was identified in a subset of patients (22 out of 39, 56.4%), with 13 patients in the New RHD group and 9 in the Known RHD group. The severity of MS also varied: 9 patients (23.1%) had mild MS, 6 patients (15.4%) had moderate MS, and 7 patients (17.9%) had severe MS. In the New RHD group, 4 patients had mild MS, 5 had moderate MS, and 4 had severe MS; whereas in the Known RHD group, 5 patients had mild MS, 1 had moderate MS, and 3 had severe MS. Twenty patients (51.3%) had both MR and MS concurrently, while 2 patients (5.1%) had isolated MS without MR. Both cases of isolated MS were in the New RHD group. (Table 9)

**Table 9:** Pattern of Mitral valve involvement among children with RHD admitted to HFCSH.

| Valve lesion | RHD type |  |  |
| --- | --- | --- | --- |
|  | New (N=25) | Known (N=14) | Total (N=39) |
| **Mitral Regurgitation (MR)** |  |  |  |
| No | 2 (8) | 0 (0) | 2 (5.13) |
| Mild | 2 (8) | 1 (7.14) | 3 (7.69) |
| Moderate | 5 (20) | 2 (14.28) | 7 (17.95) |
| Severe | 16 (64) | 11(78.57) | 27 (69.23) |
| **Mitral Stenosis (MS)** | **New (N=25)** | **Known (N=14)** | **Total (N=39)** |
| No | 12 | 5 | 17 (43.59) |
| Mild | 4 | 5 | 9 (23.08) |
| Moderate | 5 | 1 | 6 (15.38) |
| Severe | 4 | 3 | 7 (17.95) |

A substantial portion of patients (26 out of 39, or 66.7%) had aortic valve involvement. The severity of aortic regurgitation (AR) varied: fourteen patients (35.9%) had mild AR, 11 patients (28.2%) had moderate AR, and only one patient (2.6%) had severe AR. When classified into the New and Known RHD groups, in the New RHD group of 25, 9 had mild AR, 7 had moderate AR, and none had severe AR; whereas among the Known RHD group of 14, 5 had mild AR, 4 had moderate AR, and 1 had severe AR. Only one patient (2.6%) had aortic stenosis (AS), and it was severe and present in the New RHD group. This severe AS co-existed with MR. (Table 10)

**Table 10:**
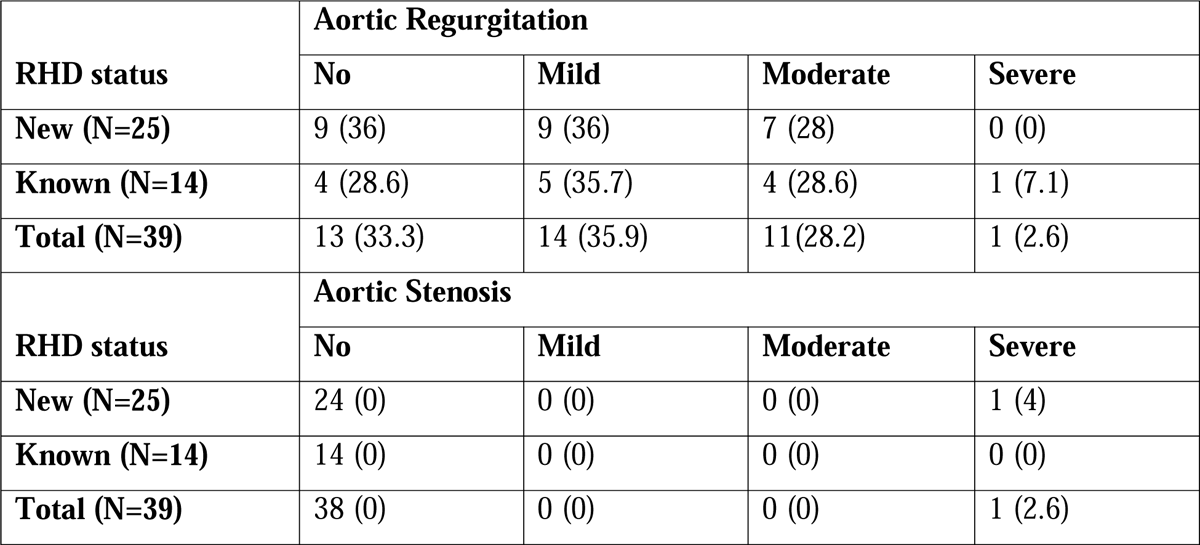
Pattern of aortic valve involvement among children with RHD in HFCSH.

Pulmonic valve involvement was less frequent than involvement of the mitral and aortic valves. Among 39 patients, 7 (17.9%) had pulmonic regurgitation, with 4 from the new RHD group and 3 from the known RHD group. No patients had pulmonic stenosis. (Table 11)

**Table 11:**
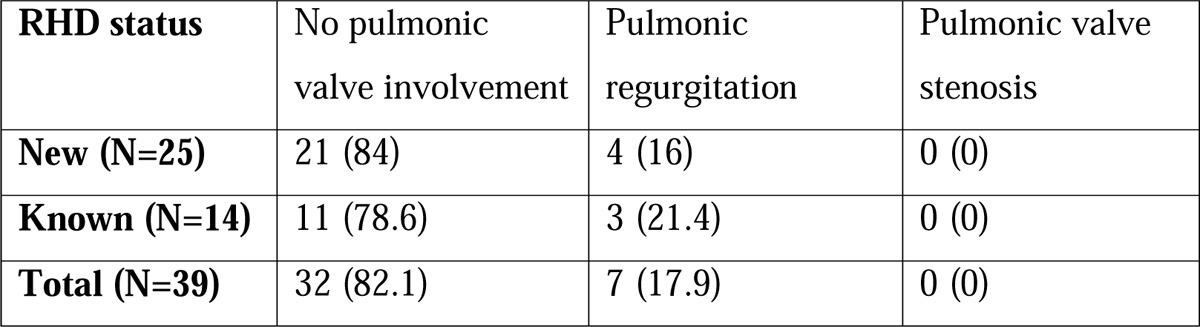
Pattern of pulmonic valve involvement among children with RHD in HFCSH.

| **RHD status** | No pulmonic valve involvement | Pulmonic regurgitation | Pulmonic valve stenosis |
| --- | --- | --- | --- |
| **New (N=25)** | 21 (84) | 4 (16) | 0 (0) |
| **Known (N=14)** | 11 (78.6) | 3 (21.4) | 0 (0) |
| **Total (N=39)** | 32 (82.1) | 7 (17.9) | 0 (0) |

**Table 12:**
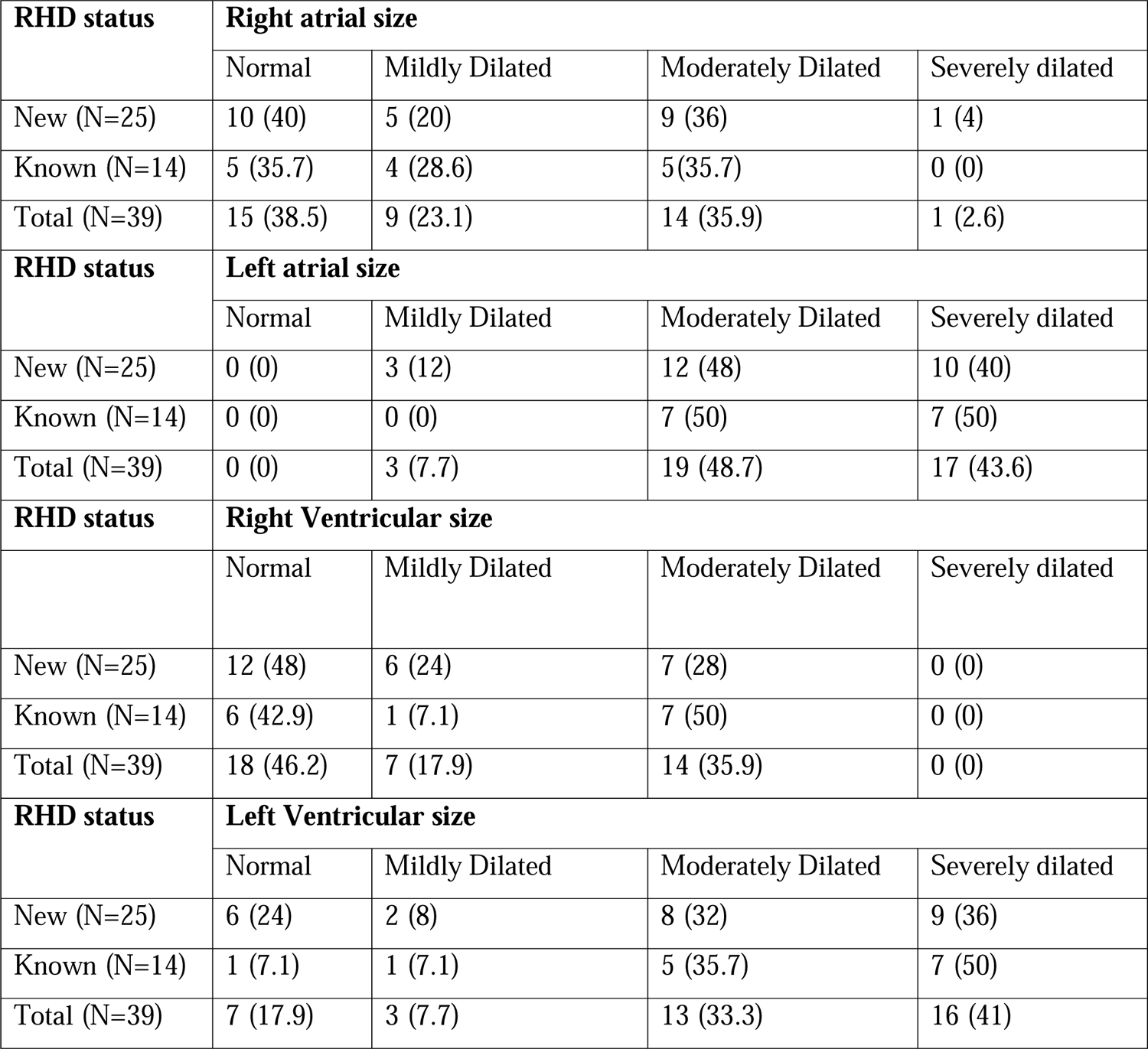
Pattern of cardiac chamber involvement among children with RHD in HFCSH.

| RHD status | Right atrial size |  |  |  |
| --- | --- | --- | --- | --- |
|  | Normal | Mildly Dilated | Moderately Dilated | Severely dilated |
| New (N=25) | 10 (40) | 5 (20) | 9 (36) | 1 (4) |
| Known (N=14) | 5 (35.7) | 4 (28.6) | 5(35.7) | 0 (0) |
| Total (N=39) | 15 (38.5) | 9 (23.1) | 14 (35.9) | 1 (2.6) |
| RHD status | Left atrial size |  |  |  |
|  | Normal | Mildly Dilated | Moderately Dilated | Severely dilated |
| New (N=25) | 0 (0) | 3 (12) | 12 (48) | 10 (40) |
| Known (N=14) | 0 (0) | 0 (0) | 7 (50) | 7 (50) |
| Total (N=39) | 0 (0) | 3 (7.7) | 19 (48.7) | 17 (43.6) |
| RHD status | Right Ventricular size |  |  |  |
|  | Normal | Mildly Dilated | Moderately Dilated | Severely dilated |
| New (N=25) | 12 (48) | 6 (24) | 7 (28) | 0 (0) |
| Known (N=14) | 6 (42.9) | 1 (7.1) | 7 (50) | 0 (0) |
| Total (N=39) | 18 (46.2) | 7 (17.9) | 14 (35.9) | 0 (0) |
| RHD status | Left Ventricular size |  |  |  |
|  | Normal | Mildly Dilated | Moderately Dilated | Severely dilated |
| New (N=25) | 6 (24) | 2 (8) | 8 (32) | 9 (36) |
| Known (N=14) | 1 (7.1) | 1 (7.1) | 5 (35.7) | 7 (50) |
| Total (N=39) | 7 (17.9) | 3 (7.7) | 13 (33.3) | 16 (41) |

**Table 02:** RHD Status vs Heart Failure.

| RHD Status | Class IV heart failure |  |  |  |
| --- | --- | --- | --- | --- |
|  | Yes |  | No | Total |
| New | 23 |  | 2 | 25 |
| Known | 12 |  | 2 | 14 |
| Total | 35 |  | 4 | 39 |
| Fisher's exact: 0.455; Cramér's V = 0.0994 |  |  |  |  |
| RHD Status | Comorbidities |  |  |  |
|  | No | Severe Acute Malnutrition | Severe Anemia | Total |
| New | 18 | 3 | 4 | 25 |
| Known | 11 | 2 | 1 | 14 |
| Total | 29 | 5 | 5 | 39 |
| Fisher's exact: P= **0.867**; Cramér's V = **0.1279** |  |  |  |  |
| RHD status | Presenting Complaint |  |  |  |
|  | Cough | Shortness of breath | Other | Total |
| New | 10 | 12 | 3 | 25 |
| Known | 5 | 9 | 0 | 14 |
| Total | 15 | 21 | 3 | 39 |
| Fisher's exact: P= 0.477; Cramér's V = 0.2356 |  |  |  |  |
| RHD Status | Rheumatic Fever |  |  |  |
|  | Yes |  | No | Total |
| New | 1 |  | 24 | 25 |
| Known | 3 |  | 11 | 14 |
| Total | 4 |  | 35 | 39 |
|  | Fisher's exact: **0.123**; Cramér's V = **-0.2756** |  |  |  |

| **RHD Status** | **Infective endocarditis** |  |  |  |
| --- | --- | --- | --- | --- |
|  | No | Possible | Definite | Total |
| **New** | 23 | 1 | 1 | 25 |
| **Known** | 11 | 2 | 1 | 14 |
| **Total** | 34 | 3 | 2 | 39 |
|  | Fisher's exact: **0.474**; Cramér's V = **0.2021** |  |  |  |

**Table 13:** Pattern of valve lesions versus RHD status.

| RHD status | Mitral Regurgitation |  |  |  |  |
| --- | --- | --- | --- | --- | --- |
|  | No | Mild | Moderate | Severe | Total |
| New | 2 | 2 | 5 | 16 | 25 |
| Known | 0 | 1 | 2 | 11 | 14 |
| Total | 2 | 3 | 7 | 27 | 39 |
|  | Fisher's exact: P= 0.806; Cramér's V = 0.2005 |  |  |  |  |
| RHD status | Mitral Stenosis |  |  |  |  |
|  | No | Mild | Moderate | Severe | Total |
| New | 12 | 4 | 5 | 4 | 25 |
| Known | 5 | 5 | 1 | 3 | 14 |
| Total | 17 | 9 | 6 | 7 | 39 |
|  | Fisher's exact: P= 0.466; Cramér's V = 0.2743 |  |  |  |  |
| RHD status | Aortic Regurgitation |  |  |  |  |
|  | No | Mild | Moderate | Severe | Total |
| New | 9 | 9 | 7 | 0 | 25 |
| Known | 4 | 5 | 4 | 1 | 14 |
| Total | 13 | 14 | 11 | 1 | 39 |
|  | Fisher's exact: P= 0.696; Cramér's V = 0.2228 |  |  |  |  |
| RHD status | Aortic Stenosis |  |  |  |  |
|  | No | Mild | Moderate | Severe | Total |
| New | 24 | 0 | 0 | 1 | 25 |
| Known | 14 | 0 | 0 | 0 | 14 |
| Total | 38 | 0 | 0 | 0 | 39 |
|  | Fisher's exact: P= 0.641; Cramér's V = -0.1214 |  |  |  |  |
| RHD status | Pulmonic valve involvement |  |  |  | Total |
|  | No |  | Yes |  |  |
| New | 21 |  | 4 |  | 25 |
| Known | 11 |  | 3 |  | 14 |
| Total | 32 |  | 7 |  | 39 |
|  | Fisher's exact: P= 0.493; Cramér's V = 0.0679 |  |  |  |  |

The pressure gradient measured across the tricuspid valve averaged 27.17 mmHg, with standard deviation of 19.734 mmHg. The right atrial (RA) size varied among patients. A normal RA size was observed in nearly 40% (15 patients), while mild, moderate, and severe dilation were identified in 21%, 37%, and 2.6% of patients, respectively.

When analyzed by subgroup, 10 patients in the new RHD group and 5 in the known RHD group had a normal RA size. Conversely, 5 patients in the new group and 4 in the known group had mildly dilated RA, 9 in the new group and 5 in the known group had moderately dilated RA, and 1 patient in the new group had severely dilated RA.

Similarly, right ventricular (RV) size varied, with normal dimensions in almost half (47%) of patients. Mild and moderate dilation were seen in 18% and 34% of patients, respectively. Among the 25 new RHD patients, 12 had a normal RV size, 6 had mildly dilated RV, and 7 had moderately dilated RV. Among the 14 known RHD cases, 6 had a normal RV size, 1 had mildly dilated RV, and 7 had moderately dilated RV.

All 39 RHD patients had a dilated left atrium. Among these, 3 patients (7.7%), all from the new RHD group, had a mildly dilated left atrium. Nineteen patients (48.7%), including 12 from the new group and 7 from the known group, had a moderately dilated left atrium. Seventeen patients (43.6%), 10 from the new group and 7 from the known group, had a severely dilated left atrium.

Eighty-two percent (32 patients) had a dilated left ventricle, with 19 from the new RHD group and 13 from the known RHD group. In contrast, 17.9% (7 patients) had a normal left ventricle, with 6 from the new RHD group and 1 from the known RHD group. Additionally, 3 patients (7.7%), 2 from the new RHD group and 1 from the known RHD group, had a mildly dilated left ventricle. Thirteen patients (33.3%), 8 from the new RHD group and 5 from the known RHD group, had a moderately dilated left ventricle. Sixteen patients (41%), 9 from the new RHD group and 7 from the known RHD group, had a severely dilated left ventricle.

One important finding was the presence of a thrombus in a single patient (2.56%). Notably, left ventricular function remained normal in all cases, despite the observed chamber size variations.

### Association of RHD status with dependent variables

We conducted Fisher’s Exact Test to examine the association between RHD status and various dependent variables, including presenting symptoms, presence of ADHF, infective endocarditis, rheumatic fever, and the pattern and severity of valve lesions. A p-value and Cramér’s V were calculated. The tests did not show a significant association. (Tables)

## Discussion

This study investigated the clinical characteristics and valve lesion patterns of rheumatic heart disease (RHD) in a resource-limited setting in Eastern Ethiopia. Among the 39 children evaluated, 72% were female and 28% were male, with a median age of 10 years. These demographics are consistent with studies from Timor-Leste, which reported a median age of 11 years, India, where the mean age was 9.6 years, and another study in Ethiopia, which reported a mean age of 10.8 years [11–13]. However, the proportion of female patients in our study was higher compared to Timor-Leste (48% female), India (male-to-female ratio of 1.15:1), and another study in Ethiopia (59.9% female patients) [11–13]. These differences could be attributed to social and geographic variations and the small sample size.

Of the children evaluated, 64% were newly diagnosed with RHD, while 36% were known RHD patients. In Harare, Zimbabwe, a similar distribution was observed, with 63.6% of children newly diagnosed [14].

Shortness of breath was the most common presenting complaint in both groups. Only 14% of known RHD patients were adherent to secondary prophylaxis, lower than the 42.18% adherence rate reported in India[13].Additionally, 21% of known RHD cases had a history of rheumatic fever, compared to only 4% of newly diagnosed cases, highlighting a gap in managing RHD that might be due to a lack of awareness or clear guidelines, leading to disease progression and recurrent rheumatic fever. Fisher’s exact test showed no significant association between RHD status and occurrence of rheumatic fever, possibly due to the small sample size..

Severe acute malnutrition and severe anemia were common comorbidities in both RHD groups, consistent with a tertiary hospital study in Ethiopia that found malnutrition in 50.7% of patients [12]. At admission, 89.7% of patients had class IV heart failure, with comparable figures between the two groups. Furthermore, 62% of patients had chest examination findings suggestive of congestion, indicating congestive heart failure, again consistent with studies in Ethiopia and other low-resource settings where children with RHD often present at advanced stages [6, 9, 15–17]. However, the proportion of class IV heart failure in our study was significantly higher than reported in other Ethiopian studies (13.6%) and studies from Timor-Leste (78% for all heart failure classes combined) and India (36.4% for all heart failure classes combined) [11–13]. This suggests delayed detection and management of RHD.

Pneumonia, rheumatic fever, and infective endocarditis were common precipitating factors for heart failure in both RHD groups, similar to findings from India and Ethiopia [12, 13]. Emphasis should be placed on preventing these conditions through preventive and therapeutic approaches. Fisher’s exact test found no significant differences in presenting complaints, comorbidities, occurrence of class IV heart failure, or chest findings between the two groups, likely due to the small sample size.

The mitral valve was the most commonly affected, with mitral regurgitation occurring in 95% of patients—92% in the newly diagnosed group and 100% in the known RHD group. Severe mitral regurgitation was present in 69% of cases. Aortic regurgitation was the second most common valve lesion, occurring in 67% of patients.

Additionally, 56% had mitral stenosis, and 51% had both mitral stenosis and mitral regurgitation. The patterns and severity of valve lesions were comparable between newly diagnosed and known RHD groups, with no significant associations found using Fisher’s exact test, likely due to the small sample size.

The predominance of mitral and aortic regurgitation aligns with existing literature, underscoring the extensive valvular damage associated with pediatric RHD [9, 16, 18]. Similar findings were reported in Timor-Leste, where 98% of patients had mitral regurgitation and 67% had severe cases, with 77% also having aortic regurgitation [11]. In India, mitral regurgitation was the most common valvular lesion, while in Addis Ababa, Ethiopia, 97.1% of patients had mitral regurgitation with 62.5% being severe [12, 13]. However, the frequency of mitral stenosis in our study was higher than in Addis Ababa (31.4%) and Timor-Leste (20%) [11, 13]. This indicates advanced disease progression at presentation in our cohort.

Left heart chambers were more commonly affected, with all patients showing left atrium dilatation and 82% having dilated left ventricles. Moderate to severe left atrial dilatation was present in 92% of RHD patients, and 74% had moderate to severe left ventricular dilatation. Additionally, 61% of patients had right atrium dilatation, and 53% had right ventricular dilatation. These patterns and severities of chamber dilatation were similar between newly diagnosed and known RHD groups, with no significant associations found using Fisher’s exact test. Our findings show a higher prevalence of chamber dilatation compared to Western and Central Africa, where left ventricular dilatation was present in 13.6% of patients and left atrial dilatation in 13.8% [19]. This further supports the notion of delayed diagnosis and advanced disease at presentation in our cohort.

### Limitations

Several limitations should be considered when interpreting the findings of this study. Firstly, the relatively small sample size and single-center design may limit the generalizability of our results to other settings within Ethiopia and beyond. Furthermore, the lack of long-term follow-up data precludes a comprehensive assessment of treatment outcomes and disease progression in this population. Future studies with larger sample sizes, prospective designs, and longer follow-up periods are needed to address these limitations and provide a more robust understanding of RHD epidemiology and outcomes in Eastern Ethiopia.

## Conclusion

This study examined the clinical characteristics of RHD in children from Eastern Ethiopia, comparing newly diagnosed and known RHD patients. A significant proportion (64%) were newly diagnosed. Shortness of breath was the primary complaint, indicating advanced disease, despite a similar age range to studies from other resource-limited settings. Comparing newly diagnosed and known RHD patients revealed no significant differences in presenting complaints, but a trend towards more severe valve lesions and a history of rheumatic fever in the known RHD group. This, coupled with low adherence to secondary prophylaxis (14%), suggests potential disease progression due to delayed diagnosis. Both groups presented with high rates of severe malnutrition and anemia, potentially contributing to disease severity. Notably, a significantly higher proportion of patients in our study had advanced heart failure (class IV), higher rates of mitral stenosis, and more chamber dilatation compared to other reports, highlighting a concerning pattern of delayed diagnosis and management.

The high prevalence of newly diagnosed RHD cases and the concerning rates of severe heart failure highlight the critical public health challenge posed by RHD in Eastern Ethiopia. This study underscores the need for improved primary healthcare infrastructure to facilitate early detection and management of RHD. Additionally, strengthening rheumatic fever prevention programs and developing effective strategies for secondary prophylaxis adherence are crucial to preventing disease progression and improving long-term outcomes for children with RHD.

### Data Sharing Statement

Any of the data used for analysis in the study is available from the corresponding author and ready to be provided up on reasonable request.

### Contributors

Conceptualization: Temesgen T Libe, Yunus E Kelil, Samrawit Abebaw, Faisel, and Kibrom Mesfin; formal analysis: Temesgen T Libe, Faisel, and Yunus E Kelil; investigation: Temesgen T Libe, Faisel, Yunus E Kelil, Samrawit Abebaw, and Kibrom; methodology: Temesgen T Libe, Faisel, and Kibrom; Validation: Temesgen T Libe, Faisel, and Kibrom, Yunus E Kelil and Samrawit Abebaw; writing the original draft: Temesgen T Libe,and Samrawit Abebaw.

## Data Availability

All data produced in the present study are available upon reasonable request to the authors

## Acknowledgments

We are very grateful to the Haramaya University College of Health and Medical Sciences for its ethical clearance provision and financial support. We also acknowledge our study participants, data collectors and supervisors for enabling us to realize this research work.

